# Phenotypical predictors of Restless Legs Syndrome in pregnancy and their association with basal ganglia and the limbic circuits

**DOI:** 10.1101/2020.07.21.20158634

**Authors:** Natalia Chechko, Jeremy Lefort-Besnard, Tamme W. Goecke, Markus Frensch, Patricia Schnakenberg, Susanne Stickel, Danilo Bzdok

## Abstract

The pregnancy-related restless legs syndrome (RLS) is thought to have a multifactorial etiology. However, the reason behind the manifestation of RLS during pregnancy remains largely elusive.

A neurological and obstetrical cohort of 308 postpartum women was screened for RLS symptoms twice: 1 to 6 days (T0) and 12 weeks postpartum (T1). 57 participants were identified as affected by pregnancy-associated RLS. The clinical and anamnestic indicators of the condition were assessed by a pattern-learning classifier trained to predict the RLS status. Structural MRI was obtained from 25 of the 57 participants with RLS history in pregnancy. In this sample, a multivariate two-window algorithm was employed to systematically chart the relationship between brain structures and phenotypical predictors.

The RLS prevalence rate in our sample was 19% (n=57), with the women suffering from RLS being older, more often unmarried, affected by gestational diabetes and having been more exposed to stressful life events. A history of RLS and the severity and frequency of repetitive compulsive movements were found to be the strongest predictors of RLS manifestation. In the RLS group, high cortisol levels, being married and receiving iron supplements were found to be associated with increased volumes in the bilateral striatum.

Investigating pregnancy-related RLS in a frame of brain phenotype modes may help shed light on the heterogeneity of the condition.

## INTRODUCTION

Restless legs syndrome (RLS) is a common sensorimotor disorder with a 5-10% prevalence in the adult population (Ohayon et al., 2012). Clinically, this condition is characterized by 1) an urge, triggered by unpleasant sensations, to move the legs, 2) momentary relief following movement, 3) worsening of the sensations with rest, and 4) a tendency for the sensations to occur in the evening or at night.

With two recognized forms, primary (idiopathic) and secondary, RLS (in both of its forms) is considered to be a continuous spectrum with a genetic contribution at one end and an environmental or comorbid disease contribution at the other (Trenkwalder et al., 2016).While idiopathic (primary) RLS is thought to be strongly influenced by genetic predisposition (Winkelmann et al., 2007), the secondary form of the condition is linked to disorders such as neurodegenerative disorder, peripheral neuronal diseases, metabolic disorders, iron deficiency, anemia and diabetes (Trenkwalder et al., 2016). Another common condition associated with RLS is pregnancy.

Largely a transitory condition, pregnancy-related RLS tends to increase from 0% before pregnancy to 23% in the third trimester (Gupta et al., 2016; Srivanitchapoom et al., 2014). Experience of RLS in a previous pregnancy and family history of RLS are strong predictors of the manifestation of pregnancy-related RLS symptoms during the ongoing pregnancy (Cesnik et al., 2010). Along with the genetic predisposition, physiological adaptations during pregnancy contribute crucially to the manifestation of RLS (Srivanitchapoom et al., 2014). Changes such as the dramatic increases in estrogen and progesterone levels (Duarte-Guterman et al., 2019), insulin resistance (Sonagra et al., 2014), hypervolemia (Duarte-Guterman et al., 2019), weight gain (Gao et al., 2009) and increased iron use (Haider et al., 2013) can render pregnant women particularly susceptible to the development of RLS symptoms.

The diagnosis of RLS in pregnancy (analogous to other forms of RLS) is based on the criteria developed by the International Restless Legs Syndrome Study Group (IRLSSG), according to which RLS is characterized by a number of neurological and other clinical indicators including the severity and frequency of repetitive compulsive movements as well as sleep and mood disturbances (Horiguchi et al., 2003). Pregnancy-related RLS is not only a neurological condition but also an obstetrical one. In particular, pregnancy-related RLS is linked to gestational diabetes mellitus (Trenkwalder et al., 2016) or pregnancy-induced hypertension (Ma et al., 2015). Thus, the manifestation of RLS in pregnancy may herald the risk of cardiovascular and metabolic diseases. In addition, multiparity (Berger et al., 2004), anemia and age (RLS is seen more frequently in older women) (Chen et al., 2012; Ohayon and Roth, 2002; Tunç et al., 2007) have been suggested to be further phenotypical predictors of pregnancy-related RLS. Finally, RLS frequently co-occurs with psychiatric disorders, particularly with affective disorders (Kallweit et al., 2016). In the context of pregnancy and childbirth, pre-pregnancy RLS has been found to be linked to perinatal depression (Wesström et al., 2014).

In sum, RLS in pregnancy is a complex sensorimotor neurological disturbance with major obstetrical and environmental contributions, causing not only physical but also substantial psychological distress (Castillo et al., 2014). To understand this condition, the complex interaction of clinical, environmental and anamnestic aspects needs to be taken into account.

In a large neurological and obstetrical sample of 308 early-postpartum women, we sought to investigate a number of possible neurological, obstetrical and anamnestic phenotypical predictors of RLS in pregnancy (e.g. history of RLS, clinical character of symptoms, age, gestational diabetes or hypertension, stressful life events (SLE), socioeconomic characteristics, multiparity, duration of pregnancy and child’s weight). Adding a brain structure analysis, we assessed the phenotypical predictors of RLS in pregnancy in a brain phenotype model. Here, an innovative multivariate pattern-learning algorithm (involving canonical correlation analysis or CCA) was employed to conjointly characterize patterns of brain structure and the phenotypical predictors of RLS, facilitating an objective assessment of their complex interplay.

## METHODS

### Study participants and clinical and neuropsychological assessments

A total of 308 women were recruited in the Department of Gynecology and Obstetrics at the University Hospital Aachen within 1 to 6 days of childbirth (T0) (Figure 1) between January 2016 and January 2018. The exclusion criteria were alcoholic or psychotropic substance dependency or use during pregnancy, anti-depressive or anti-psychotic medication during pregnancy, history of psychosis or manic episodes, depressive episode during the current pregnancy, and inadequate proficiency in German or English. To control for the exclusion criteria, a short non standardized clinical interview was conducted according to the DSM V criteria by an experienced psychiatrist (NC).

**Figure 1.**
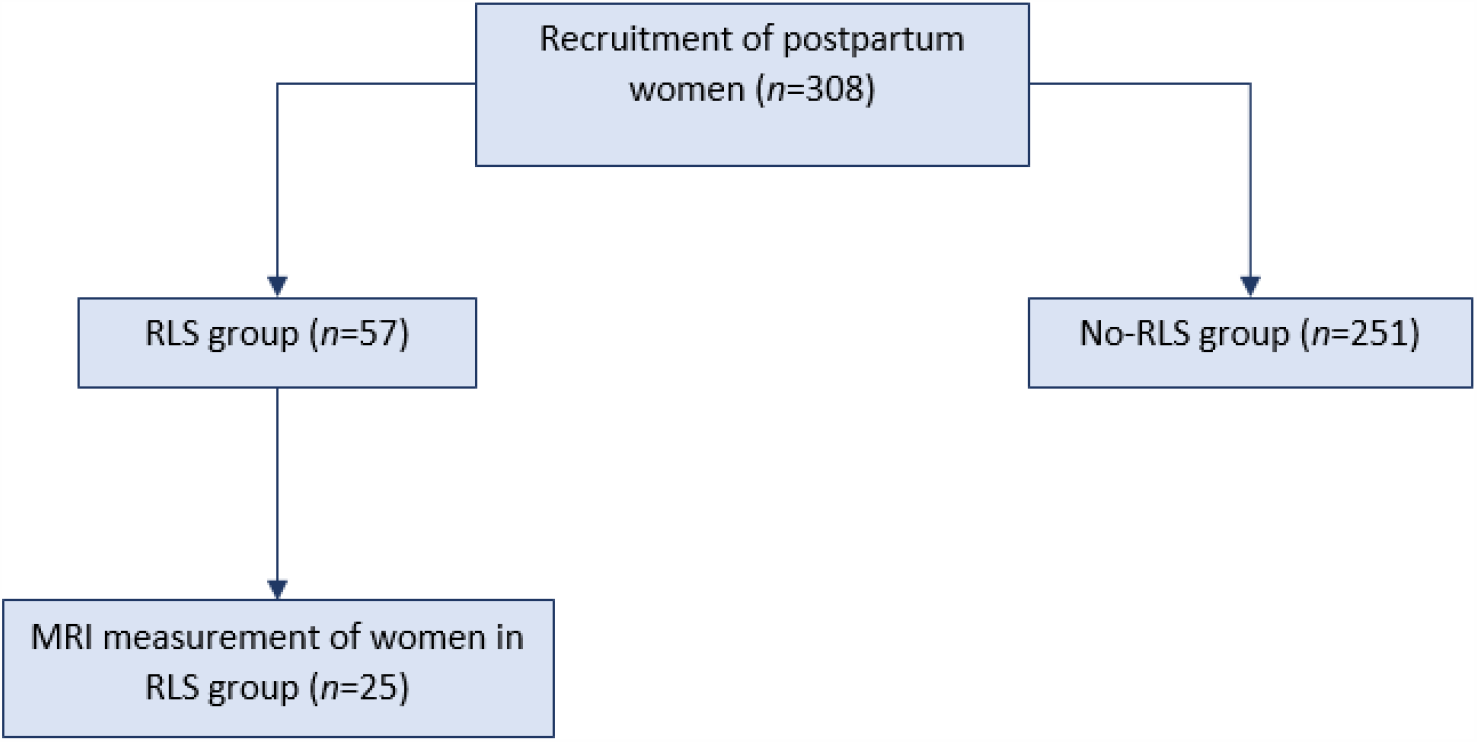
Flowchart of inclusion into the study.

Following receipt of the participants’ informed consent, all participants were screened for RLS symptoms in pregnancy based on the criteria of the International Restless Legs Syndrome Study Group (IRLSSG) (Allen et al., 2014) and by means of the Restless Legs Syndrome Rating Scale (IRLSS) (Allen et al., 2014). A detailed description of the study population is given in Tables 1A, 1B and 1C. In addition, voxel-based morphometric (VBM) measurements were collected from 25 participants with RLS history in pregnancy, aged 23-42 years, the remainder of the RLS group (32) not being eligible for the MRI. A detailed description of the sample that participated in the MRI experiment is given in Table 2 (please refer also to Figure 1 with the flowchart of inclusion into the study).

**Table 1A.**
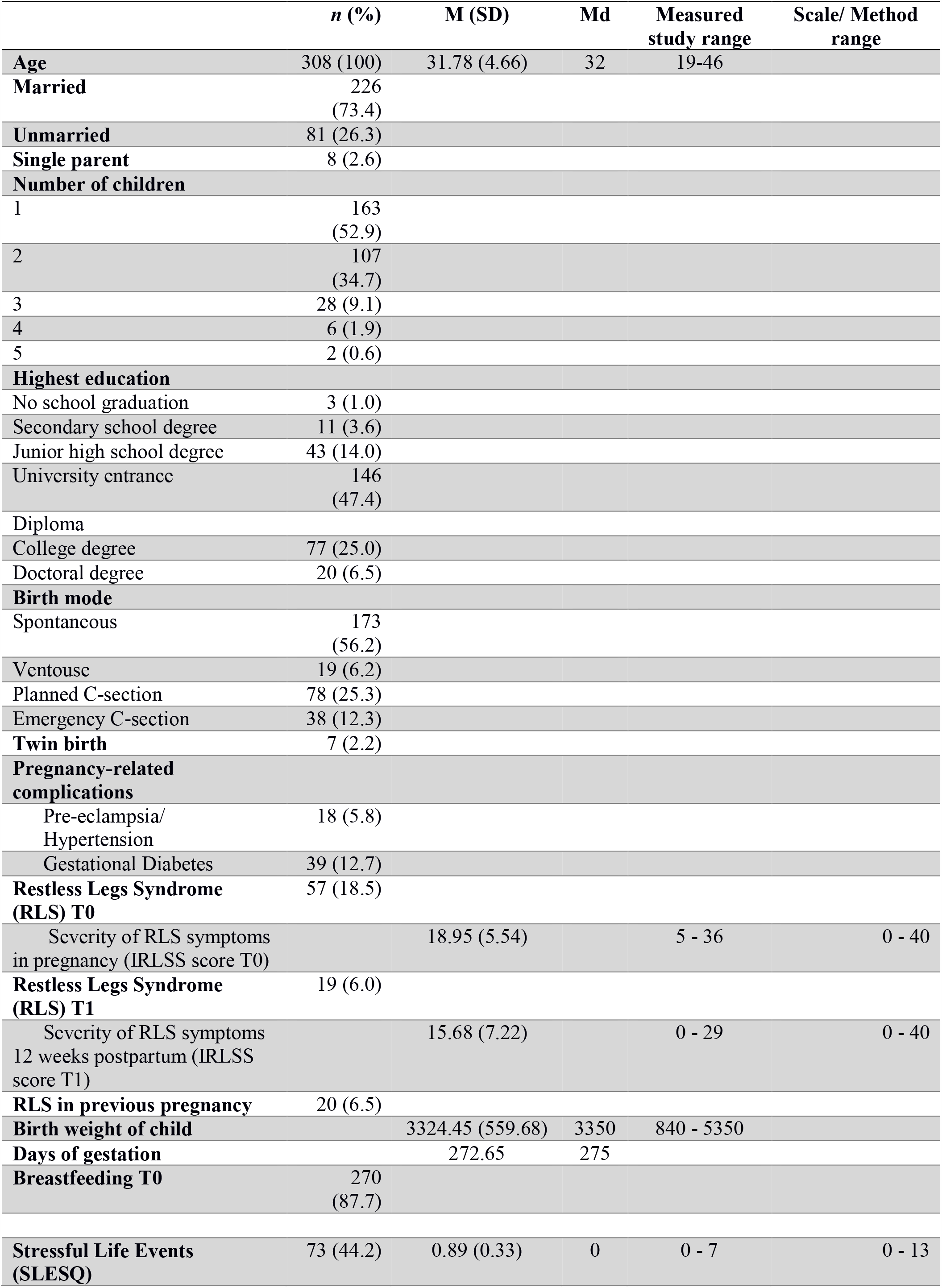
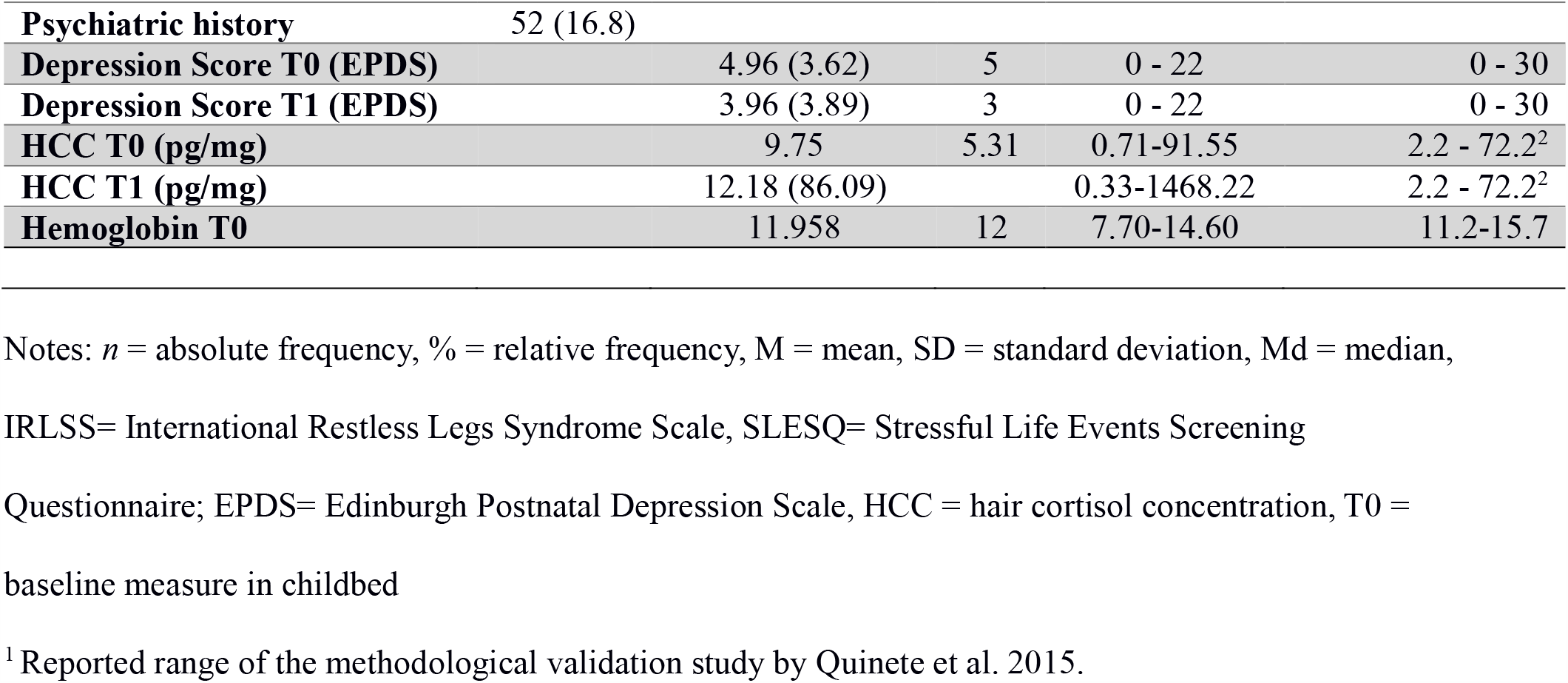
Demographics of the whole sample.

**Table 1B.** Differences between women with and without RLS.

|  | RLS<br>(*n* = 57) | No RLS<br>(*n* = 251) | *p* |
| --- | --- | --- | --- |
|  | M (SD) | M (SD) |  |
| Age | 33.18 (4.66) | 31.47 (4.608) | .012 |
| Depression score EPDS T0 | 4.72 (4.02) | 5.01 (3.53) | .583 |
| Depression score EPDS T1 | 3.77 (3.23) | 4.00 (4.03) | .690 |
| Hair Cortisol Concentration (HCC) T0 (in pg/mg) | 11.45 (16.14) | 9.36 (11.54) | .260 |
| Hair Cortisol Concentration (HCC) T1 (in pg/mg) | 5.97 (5.95) | 13.65 (95.70) | .549 |
| Hemoglobin in g/dL T0 | 12.06 (1.13) | 11.94 (1.18) | .483 |
| Length of pregnancy in days | 272.65 (9.96) | 272.66 (13.50) | .997 |
| Child's birth weight (in gram) | 3315.69 (501.96) | 3326.28 (571.94) | .902 |
|  | *n* (%) | *n* (%) |  |
| Family Status |  |  | .047 |
| Married | 36 (63.2) | 190 (76) |  |
| Unmarried | 21 (36.8) | 60 (24) |  |
| Number of children |  |  | .186 |
| 1 | 26 (45.6) | 137 (55) |  |
| 2 | 21 (36.8) | 86 (34.5) |  |
| 3 | 6 (10.5) | 22 (8.8) |  |
| 4 | 3 (5.3) | 3 (1.2) |  |
| 5 | 1 (1.8) | 1 (0.4) |  |
| Highest education |  |  | .511 |
| No school graduation | 1 (1.8) | 2 (0.8) |  |
| Secondary school degree | 3 (5.3) | 8 (3.3) |  |
| Junior high school degree | 5 (8.8) | 38 (15.6) |  |
| University entrance | 26 (45.6) | 120 (49.4) |  |
| Diploma |  |  |  |
| College degree | 16 (28.1) | 61 (25.1) |  |
| Doctoral degree | 6 (10.5) | 14 (5.8) |  |
| Birth mode |  |  | .808 |
| Spontaneous | 31 (54.4) | 142 (56.6) |  |
| Ventouse | 4 (7) | 15 (6) |  |
| Section | 13 (22.8) | 65 (25.9) |  |
| Emergency section | 9 (15.8) | 29 (11.6) |  |
| Breastfeeding at T0 | 49 (86) | 221 (88) | .666 |
| Pregnancy-related complications |  |  |  |
| Pre-eclampsia / Hypertension | 4 (7.0) | 13 (5.2) | .583 |
| Gestational Diabetes | 11 (19.3) | 29 (11.6) | .049 |
| Child relocation after birth | 69 (27.6) | 17 (29.8) | .603 |
| Iron supplements | 32 (58.2) | - |  |
| RLS in previous pregnancy | 20 (35.0) | - |  |
| Most severe RLS symptoms |  |  |  |
| First trimester | 2 (3.51) | - |  |
| Second trimester | 7 (12.28) | - |  |
| Third trimester | 38 (66.67) | - |  |
| Equally high over all trimesters | 8 (14.04) | - |  |
| Psychiatric history | 11 (19.3) | 41 (16.4) | .599 |
| Baby blues | 30 (52.6) | 108 (43.7) | .223 |
| Stressful life events (SLESQ) | 35 (61.4) | 103 (41.4) | .006 |

Notes. *n* = absolute frequency, M = mean, SD = standard deviation

**Table 1C.**
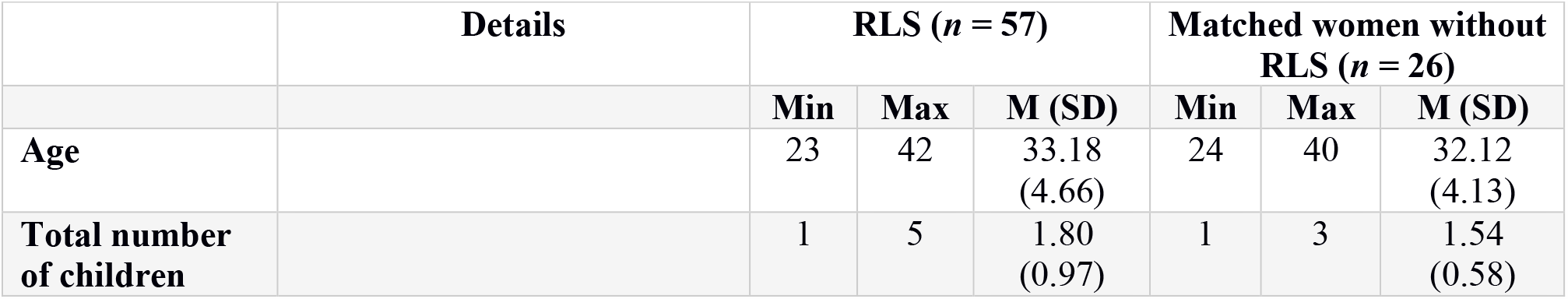
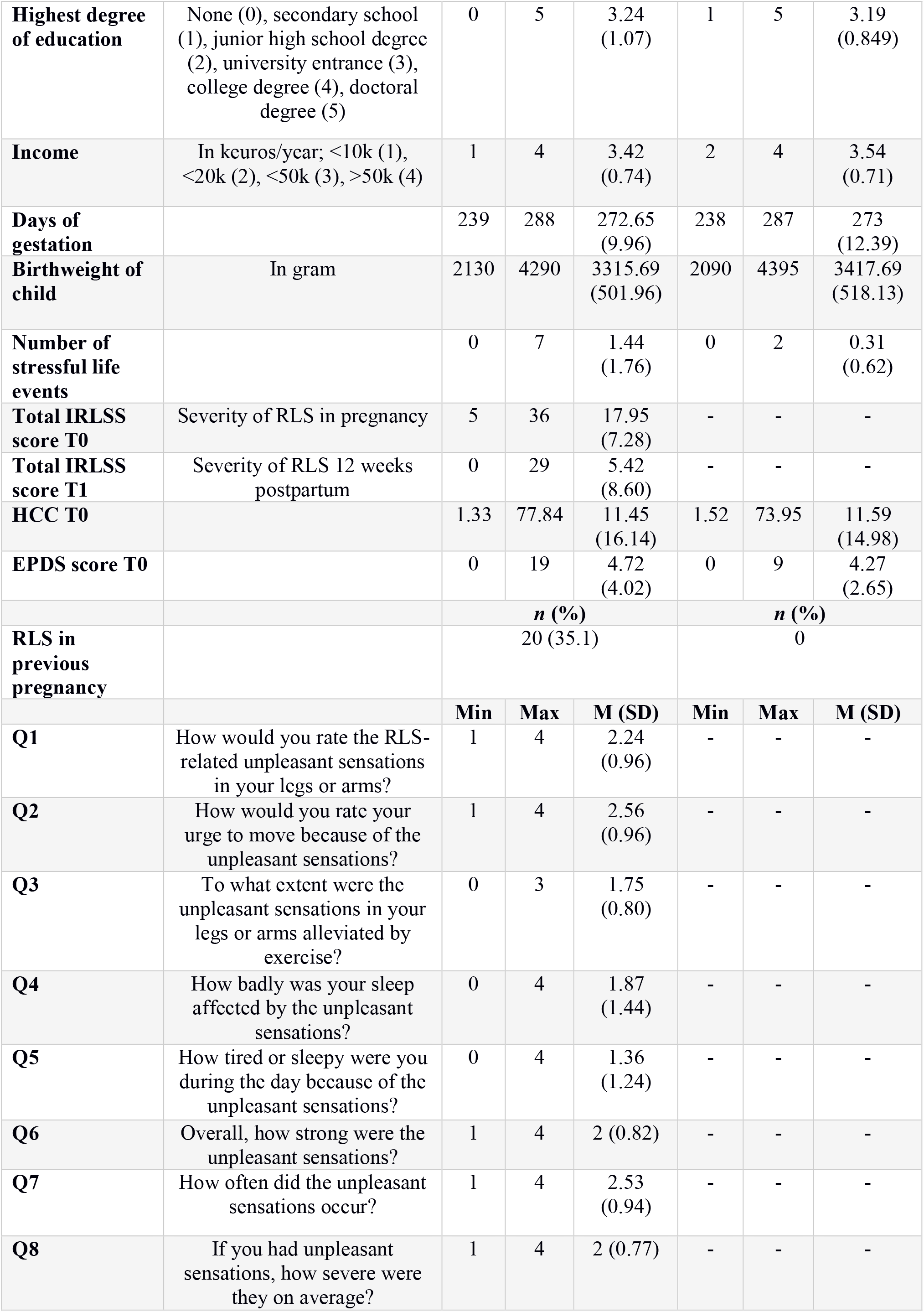
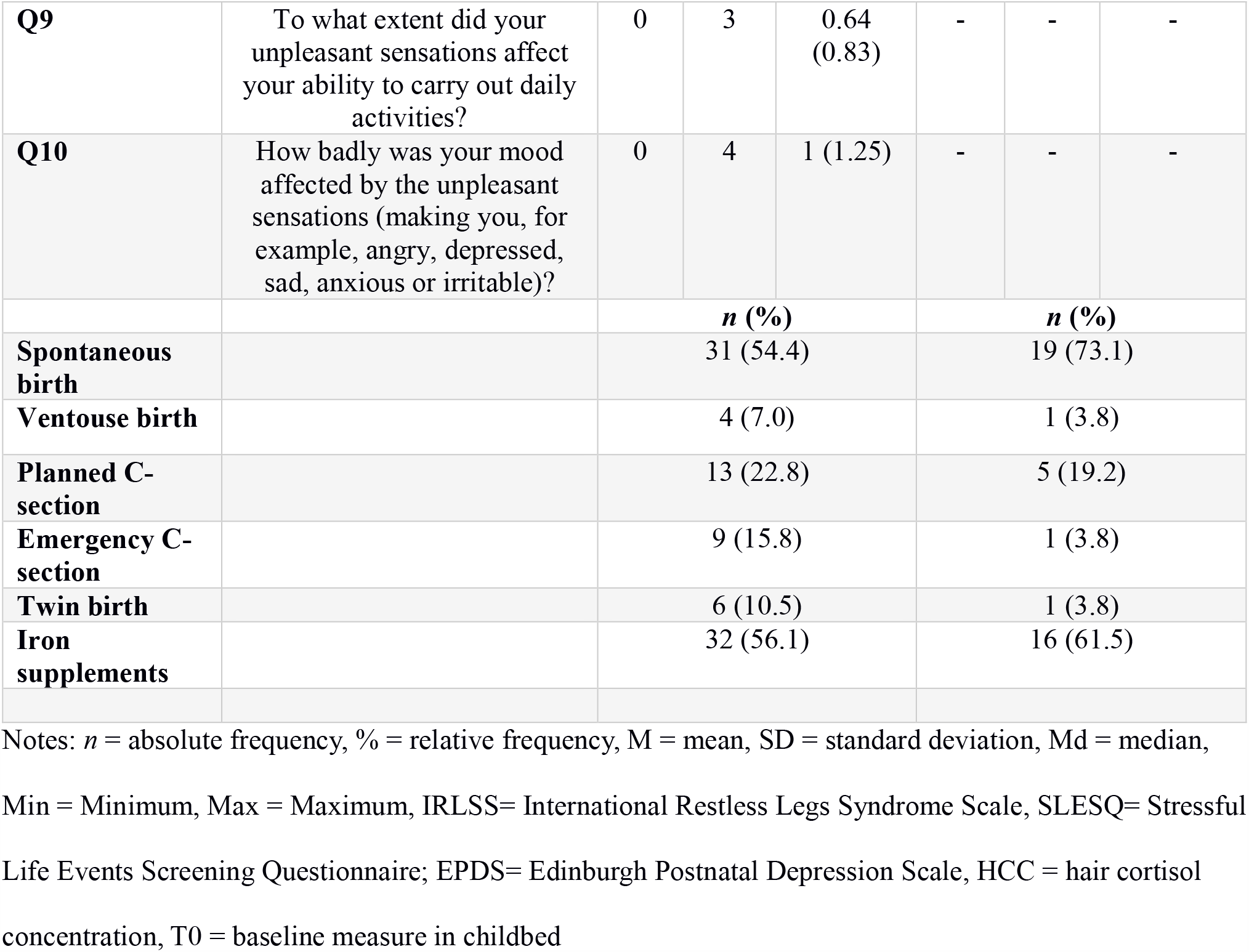
**Demographics of women with RLS and their matched counterparts, in accordance with Figure 1**

**Table 2.** **Demographics of women with RLS participated on MRI, in accordance with Figures 2 and 3**

| Variables | Descriptive statistics |  |  | Details |
| --- | --- | --- | --- | --- |
|  | Min | Max | Mean (SD) |  |
| Age | 23 | 42 | 33.08 (5.84) |  |
| Total number of children | 1 | 5 | 2.04 (1.23) |  |
|  | Min | Max | Mode |  |
| Highest degree of education | 0 | 5 | 3 | None (0), secondary school (1), junior high school degree (2), university entrance (3), college degree (4), doctoral degree (5) |
| Income | 1 | 4 | 3 | In euros/year; <10k (1), <20k (2), <50k (3), >50k (4) |
|  | Min | Max | Mean (SD) |  |
| Days of gestation | 239 | 288 | 274 (10.97) |  |
| Birth weight of child | 2700 | 6300 | 3704 (847.77) | In gram |
| Number of stressful life events | 0 | 6 | 1.2 (1.56) |  |
| IRLSS score T0 | 7 | 28 | 16.87 (6.26) | Severity of RLS symptoms in pregnancy |
| IRLSS score T1 | 0 | 29 | 7.2 (9.82) | Severity of RLS symptoms 12 weeks postpartum |
| HCC T0 | 1.32 | 77.84 | 9.2 (15.38) |  |
| EPDS score T0 | 0 | 11 | 4.37 (3.46) |  |
| RLS in previous pregnancy | 0 | 2 | 1.12 (0.79) |  |
| Q1. How would you rate the RLS-related unpleasant sensations in your legs or arms? | 1 | 3 | 2.08 (0.77) |  |
| Q2. How would you rate your urge to move because of the unpleasant sensations? | 1 | 4 | 2.37 (0.92) |  |
| Q3. To what extent were the unpleasant sensations in your legs or arms alleviated by exercise? | 1 | 3 | 1.71 (0.75) |  |
| Q4. How badly was your sleep affected by the unpleasant sensations? | 0 | 4 | 1.96 (1.43) |  |
| Q5. How tired or sleepy were you during the day because of the unpleasant sensations? | 0 | 4 | 1.37 (1.13) |  |
| Q6. Overall, how strong were the unpleasant sensations? | 1 | 3 | 1.96 (0.69) |  |
| Q7. How often did the unpleasant sensations occur? | 0 | 4 | 2.42 (0.93) |  |
| Q8. If you had unpleasant sensations, how severe were they on average? | 0 | 2 | 0.46<br>(0.58) |  |
| Q9. To what extent did your unpleasant sensations affect your ability to carry out daily activities? | 0 | 2 | 0.46<br>(0.58) |  |
| Q10. How badly was your mood affected by the unpleasant sensations (making you, for example, angry, depressed, sad, anxious or irritable)? | 0 | 3 | 0.71<br>(0.91) |  |
|  | **N (%)** |  |  |  |
| No birth-related complications | 17 (68.00) |  |  |  |
| Birth-related complications | 8 (32.00) |  |  |  |
| No pregnancy-related complications | 11 (44.00) |  |  |  |
| Pregnancy-related complications | 14 (56.00) |  |  |  |
| With partner, not married | 12 (48.00) |  |  |  |
| With partner, married | 13 (52.00) |  |  |  |
| No breastfeeding | 6 (24.00) |  |  |  |
| Breastfeeding | 19 (76.00) |  |  |  |
| No relocation of child to other ward | 18 (72.00) |  |  |  |
| Relocation of child to other ward | 7 (28.00) |  |  |  |
| Birth of a single child | 21 (84.00) |  |  |  |
| Twin birth | 4 (16.00) |  |  |  |
| No iron supplements | 9 (36.00) |  |  |  |
| Iron supplements | 16 (64.00) |  |  |  |

In addition to the IRLSS, the following questionnaires were used at T0: the Stressful Life Events Screening Questionnaire (SLESQ) (Goodman et al., 1998) was used as a self-report instrument to help assess the participants’ encounter with 13 particularly traumatic experiences, and the Edinburgh Postnatal Depression Scale (EPDS) (Cox et al., 1987) was used as a self-report instrument for the screening of postpartum depression. Furthermore, the participants filled out a standardized questionnaire to help obtain anamnestic and pregnancy-related information, e.g. family history of psychiatric conditions, previous psychiatric history, income, marital status, complications during pregnancy or birth mode.

The second screening session for postpartum RLS symptoms took place 12 weeks after delivery (T1) when the IRLSS and EPDS were applied for the second time.

Hair samples were collected shortly after delivery (T0) and 12 weeks postpartum (T1) to assess the cumulative cortisol exposure over the last trimester of pregnancy and three months postpartum, respectively.

The study was conducted in compliance with the Helsinki Declaration and was approved by the local ethics committee of the Medical Faculty, RWTH Aachen University, Germany.

### Hair sample collection and preparation

Hair cortisol and cortisone are widely regarded as biomarkers of chronic stress (Stalder et al., 2017). Hair samples were obtained from the posterior vertex of the head, stored in aluminum foil, and treated as described in Stalder and Kirschbaum [24]. 3 cm of hair was cut at each time point and approximately 50 mg was weighed in a polypropylene sampling tube using a microbalance. Hair samples were washed with 2-propanol, extracted with a fourfold deuterium isotope-labeled internal standard of cortisol and methanol for 24 h and then analyzed with liquid chromatography triple quadrupole mass spectrometry using an ion trap (Agilent Technologies 1200 infinity series - QTRAP 5500 ABSciex). The limits of quantification were 0.05 ng/mL or 2 pg/mg hair, respectively.

### MRI procedure and Voxel-based morphometry

The structural image was acquired with an anatomical 3D T1-weighted MPRAGE sequence (176 slices, TR = 2300 ms, TE = 1.99 ms, FoV = 256 × 256 mm^2^, flip angle = 9°, voxel resolution = 1 × 1 × 1 mm^3^). The brain tissue was segmented into gray matter, white matter and cerebrospinal fluid, the resultant adjusted volume measurements representing the amount of gray matter corrected for individual brain size. Structural MRI data were preprocessed using the Computational Anatomy Toolbox (CAT12) and SPM12 toolbox implemented in Matlab 2015b (MathWorks, Inc., Natick, MA) to derive voxel-wise gray matter volumes for each subject. The default settings of CAT12 were applied for spatial registration, normalization and segmentation of the T1-weighted structural brain images. For a precise spatial normalization into standard (MNI), the Diffeomorphic Anatomic Registration Through Exponentiated Linear algebra algorithm (DARTEL) (Klein et al., 2009) was performed. The images were segmented into gray matter, white matter, and cerebrospinal fluid, and modulated with Jacobian determinants. Finally, the modulated gray matter images were smoothed with an 8 mm isotropic FWHM Gaussian kernel. Subsequently, the association between brain structure and clinical indicators of patients experiencing RLS in pregnancy was systematically charted.

### Assessing the clinical and anamnestic indicators in predicting RLS

Independent samples t-tests (for continuous variables) and chi-square tests (for categorical variables) were used to assess differences between the RLS and control groups. Within-group differences were analyzed by means of paired samples t-tests, with IBM Statistics 25 (SPSS, Chicago, IL) being used for the analysis.

The relative importance of the clinical and anamnestic indicators for predicting RLS was analyzed using a L2-penalized logistic regression based on the information of 57 RLS patients and 26 matched healthy controls. The L2 regularization was used to reduce the chances of overfitting, which can render the models’ prediction of future observation unreliable (Okser et al., 2014). The L2-penalized logistic regression estimated the separating hyperplane (i.e., a linear function), distinguishing between patients with RLS and healthy participants. The outcome to be predicted was defined by being healthy (0) or being a patient with RLS (1). The model parameters were then fit to optimally predict RLS based on all the standardized clinical and anamnestic data.

### Brain-phenotype association across the whole-brain gray matter

The relationship between brain structure and clinical and anamnestic indicators of patients experiencing restless legs syndrome (RLS) in pregnancy was systematically charted based on the structural magnetic resonance imaging (T1-MRI) data from RLS patients in pregnancy.

### Signal extraction

Using the Automated Anatomical Labeling (AAL) ROI atlas (Tzourio-Mazoyer et al., 2002), quantitative measures of gray matter volume were extracted within the 116 macroscopic brain structures labeled in this atlas in every patient. Widely used in neuroimaging (Fransson and Marrelec, 2008; Rubinov, 2013; Yang et al., 2013), the AAL atlas is the result of an automated anatomical parcellation of the spatially normalized single-subject high-resolution scan of the brain.

For the extraction of relevant signal from the structural brain data, the total of 116 regions served as topographic masks to average the volume information across the voxels belonging to a given region. Each AAL region was represented by the average gray matter volume across all AAL region voxels. This way of engineering the morphological brain features yielded as many volumetric brain variables per patient as the total number of AAL regions (i.e., 116). All region-wise structural volumes were transformed into z-scores by mean centering and unit-variance scaling (Gelman and Hill, 2006).

### Joint multivariate decomposition across the whole brain

A multivariate pattern analysis technique, namely the canonical correlation analysis (CCA), was employed (Wang et al., 2020). This approach relates multiple blocks of data (with structural imaging features on the one hand, and anamnestic and clinical indicators on the other) through a latent factor model. Projections of the first 10 principal components of the standardized volumetric brain measures (i.e., 25 * 10 matrix) as well as the first 10 principal component projections of the standardized anamnestic/clinical data (i.e., 25 * 10 matrix) were fed into a CCA algorithm, the number 25 indicating the number of subjects included in the analysis. This multivariate and multimodality approach enabled us to objectively assess the relationship between patient differences in brain volume and anamnestic/clinical indicators. The CCA determines the canonical vectors u and v that maximize the symmetric relationship between a linear combination of brain volumes (X) and a linear combination of anamnestic and clinical data (Y), thereby identifying the two projections, Xu and Yv, that yield maximal linear co-occurrence between sets of regions with volumetric changes and sets of patients’ clinical changes. In concrete terms, the positive (negative) modulation weights revealed increased (decreased) strengths of brain-behavior association relative to the baseline volumetric changes.

In sum, using the CCA, which finds linear combinations to optimize correlations between brain region volumes and an array of anamnestic/clinical measures, we estimated pairs of canonical variates along which sets of anamnestic/clinical measures and patterns of brain volume correlated coherently across subjects. Henceforth, we refer to each pair of such variates as a ‘mode’ of coherent brain-phenotype co-variation.

### Testing for statistically significant brain-behavior associations between brain volume and phenotypical indicators

Following the CCA of RLS patients, the statistical robustness of the ensuing brain-phenotype relationship was assessed through a non-parametric permutation approach (Efron, 2012; Nichols and Holmes, 2002) using the canonical correlation as the test statistic. Relying on minimal modeling assumptions, a valid null distribution was derived for the achieved correlation between the canonical variates resulting from the CCA analysis. In 1,000 permutation iterations, the brain volume matrix was held constant, while the anamnestic and clinical data matrix underwent patient-wise random shuffling. While the constructed surrogate data preserved the statistical structure idiosyncratic to the MRI-derived signals, they were permitted to selectively destroy the signal property related to the CCA statistic to be tested (Wang et al., 2020). The empirical distribution thus generated reflected the null hypothesis of random association between volume and anamnestic and clinical indicators across patients. The Pearson correlation r between the perturbed canonical variates were recorded in each iteration, with the P-values being obtained from the number of correlations r from the null CCA model. This analysis revealed a single significant CCA mode that relates brain volume to subjects’ clinical and anamnestic measures (p < 0.05).

### Post-hoc brain-phenotype association of the basal ganglia

With the whole-brain results showing the basal ganglia to be the site of the most important and inter-hemispheric variations across brain volumes, we further investigated patterns of changes in the basal ganglia specifically related to the anamnestic and clinical indicators. Quantitative measures of gray matter volume were extracted within the basal ganglia brain structures, the left and right caudate and putamen, as labeled in AAL in every participant. This way of engineering morphological brain features yielded as many volumetric brain variables per patient as the total number of voxels included in these AAL regions (i.e., 10881). All region-wise structural volumes were transformed into z-scores by mean centering and unit-variance scaling. The first 10 principal components of the standardized basal ganglia volumetric brain measures as well as the first 10 principal components of the standardized clinical and anamnestic data were fed into a CCA. Following the preceding whole-brain CCA (cf. previous paragraph), only the first mode was automatically computed.

### Code availability

Python was selected as the scientific computing engine, the open-source ecosystem of which helps enhance replicability, reusability and provenance tracking. Scikit-learn (Pedregosa et al., 2011) provided efficient, unit-tested implementations of state-of-the-art statistical learning algorithms (http://scikit-learn.org). All analysis scripts of the present study are readily accessible to the reader online (https://github.com/JLefortBesnard/RLS_2019).

## RESULTS

### Course of RLS in pregnancy

57 (19%) of the 308 women reported to experience RLS symptoms during pregnancy. While for 22 (39%) of them this was the first pregnancy, for 35 (61%), it was a second or third pregnancy. Of these 35 women, 20 (57%) had experienced RLS symptoms during a previous pregnancy. 19 (33%) of the 57 women continued to experience RLS symptoms 12 weeks after delivery. However, the severity of RLS symptoms 12 weeks after delivery (T1) (M = 15.68, SD = 7.22) was significantly lower compared to the severity of RLS symptoms at T0 (M = 18.95, SD = 5.54), t(18) = 3.314, p = 0.04. For 2 (4%) women, no information was available with respect to RLS symptoms 12 weeks postpartum. 38 women (67%) reported the RLS symptoms to have been most severe during the third trimester of pregnancy (for details, see Table 1A).

### Differences between RLS and control groups

Participants with RLS symptoms during pregnancy were older (Mdn = 35) compared to their counterparts without RLS (Mdn = 31), U = 8.688, p = 0.11. Also, women in the RLS group had more often gestational diabetes (n = 10, 17.5%) compared to those in the control group without RLS (n = 29, 11.6%), τ^2^ (1, 308) = 6.040, p = .049. In addition, a significantly larger number of women in the RLS group (n = 35, 61.4%) experienced SLE compared to the control group (n = 103, 41.4%), τ^2^ (1, 306) = 7.522, p = .006 and were not married, τ^2^ (1, 307) = 3.941, =.047 (see Table 1B). On the other hand, the groups with and without RLS did not differ with respect to depressivity (EPDS) and HCC at any time point and Hemoglobin in g/dL at T0, and also in terms of severity of baby blues, birth mode, multiparity and psychiatric history.

### Detecting behavioral descriptors that distinguish patients with RLS

In a group of 57 RLS patients and 26 matched healthy controls, we explored the contribution of each anamnestic and clinical indicator to the detection of patients with RLS. Eight out of 28 variables were highly weighted for detecting patients with RLS. In addition to the total and IRLSS, further features indicative of RLS included experience of RLS in a previous pregnancy, and a high score with respect to the questions 1, 2, 3, 6, 7 and 8 (Fig. 1, Tab.1C). Here, the items describing the degree of RLS symptom relief by exercise (Q3), the urge to move the legs (Q2) and the average severity of the unpleasant sensations in the legs (Q8) were most indicative of patients with RLS. On the other hand, items describing the degree of sleep disturbance (Q4, Q5) as well as being affected in the daily activities (Q9) or mood disturbances (Q10) were the least indicative. Age, number of children, duration of pregnancy, birthweight or mode of birth also did not contribute to the detection of participants affected by pregnancy-related RLS.

### First CCA analysis: coherent morphological patterns across a whole brain atlas

The main objective of the whole-brain study was to identify components of the relationship between brain structures and clinical traits that described RLS patients in pregnancy. A single statistically significant CCA mode emerged in the first analysis (r = 0.993, p < 0.05), exhibiting a strong covariation of brain volume measures and diverse anamnestic and clinical indicators (Figure 2, Table 2). The other 9 of the 10 estimated modes were not significantly robust. The most important CCA mode comprised measures that varied along a positive-negative axis. As regards the anamnestic and clinical indicators, a high cortisol level, being married, and receiving iron supplements were located at the positive end of the mode. The brain regions at the positive end of the mode included the left superior temporal pole, the left inferior frontal gyrus, the basal ganglia (right and left caudate nucleus and putamen), and the right lobule 6 of the cerebellar hemisphere. Note that only the basal ganglia changes were located in both hemispheres. The most significant anamnestic and clinical changes located at the negative end of the mode included positive history of RLS in previous pregnancy, not being married and not receiving any iron supplement. Further anamnestic and clinical changes located at the negative end of the mode included complaining about the severity of RLS-related sensations (Q1), the degree of the urge to move the legs (Q2), and the number of SLE. Brain regions with high negative loadings included the right inferior parietal lobule, the left posterior cingulate gyrus, the right lobule 9, VIIIb, crus II and the left lobule 10 of the cerebellar hemisphere, the left olfactory cortex, the left superior frontal gyrus, the right postcentral gyrus, the left and right calcarine sulci and the right medial orbitofrontal cortex. In sum, our results identified several inter-hemispheric zones of activity, especially in the basal ganglia, contributing to coherent anamnestic and clinical changes. Based on this observation, we focused on this particular region in a second analysis.

**Figure 2.**
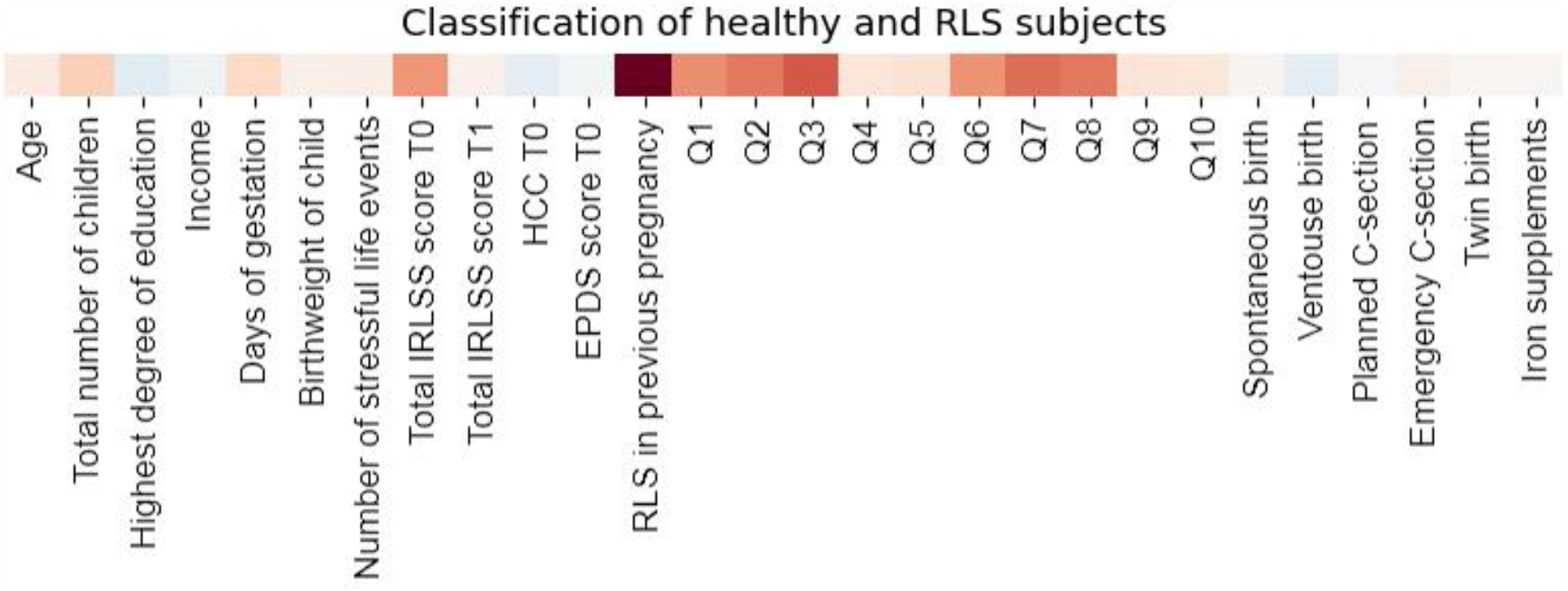
Informative features for predicting RLS. *IRLSS score T0 = Severity of RLS symptoms in pregnancy; IRLSS score T1 = Severity of RLS symptoms 12 weeks postpartum A modern constrained version of logistic regression was deployed to explore the contribution of each anamnestic and clinical indicator to the prediction of RLS experience. The x-axis depicts the variables included in the analysis. The red square indicates the feature contributing to the detection of an RLS patient while the blue square indicates the feature responsible for detecting a healthy participant. For example, a high score in the feature “RLS in previous pregnancy” would tip the balance of the output toward being an RLS patient, while a high level of cortisone (HCC T0) would tip the balance toward being a healthy participant. In sum, the most informative features with respect to the prediction of RLS were RLS in a previous pregnancy, and a high score against the questions 1, 2, 3, 6, 7 and 8.

### Second CCA analysis: exploratory voxel-level investigation within the basal ganglia

As a logical follow-up, we studied the relationship between changes in the phenotypical predictors and specific changes within the basal ganglia in RLS patients in pregnancy (Figure 3, Table 2). To that end, the first 10 principal components of standardized brain volume of the basal ganglia regions and the first 10 principal components of standardized anamnestic and clinical data were fed into the CCA, with only one CCA mode being computed given the previous results. The anamnestic and clinical changes located at the positive end of the mode included having a heavier baby and having the unpleasant sensations in the legs or arms alleviated by exercise (Q3), while the basal ganglia changes at the positive end of the mode were the left and right body and head of the caudate nucleus and the left and right inferior putamen. The amnestic and clinical changes at the negative end of the mode were being young and not being so affected by RLS on a daily basis (Q9), while the corresponding brain volume changes included the superior part of the putamen.

**Figure 3.**
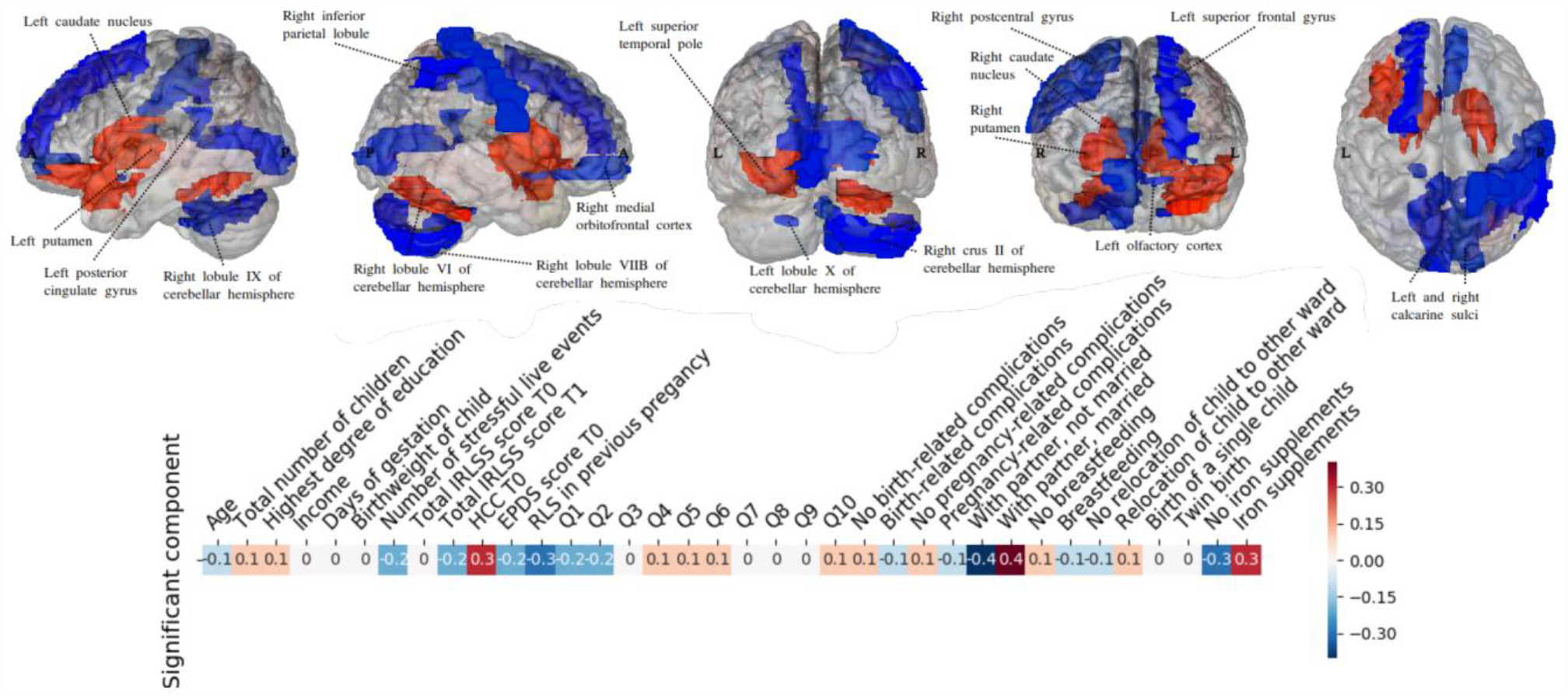
CCA reveals multivariate patterns of link between phenotypical predictors of RLS and brain structure. *IRLSS score T0 = Severity of RLS symptoms in pregnancy; IRLSS score T1 = Severity of RLS symptoms 12 weeks postpartum The figure shows the first canonical model of brain-behavior co-variation that was statistically significant based on permutation testing (p < 0.05). In the top row, the brain structure loadings are plotted from the left, right, posterior, anterior and superior views. The blue regions indicate the negative CCA loading associated with volume in this specific region, while the red region volumes indicate a positive CCA loading for this specific region. The bottom row depicts the loadings of the patient’s anamnestic and clinical indicators (blue for a negative loading and red for a positive one). A high cortisol level, being married, and receiving iron supplement were robustly linked to an increased size of the left superior temporal pole, the left inferior frontal gyrus, the basal ganglia (right and left caudal nucleus and putamen), and the right lobule 6 of the cerebellar hemisphere. On the other hand, RLS during previous pregnancy (positive history of RLS), not being married and not receiving any iron supplement were found to be linked to a decreased volume size of the right inferior parietal lobule, the left posterior cingulate gyrus, the right lobule 9, 8B, crus 2 and the left lobule 10 of the cerebellar hemisphere, the left olfactory cortex, the left superior frontal gyrus, the right postcentral gyrus, the left and right calcarine sulci and the right medial orbitofrontal cortex. Further anamnestic and clinical changes located at the negative end of the mode included complaining about the severity of RLS-related sensations (Q1), the degree of the urge to move the legs (Q2), and the number of SLE. That the most inter-hemispherically coherent brain-phenotype associations were found in the basal ganglia justified our targeted analysis (Fig. 2)

**Figure 3.**
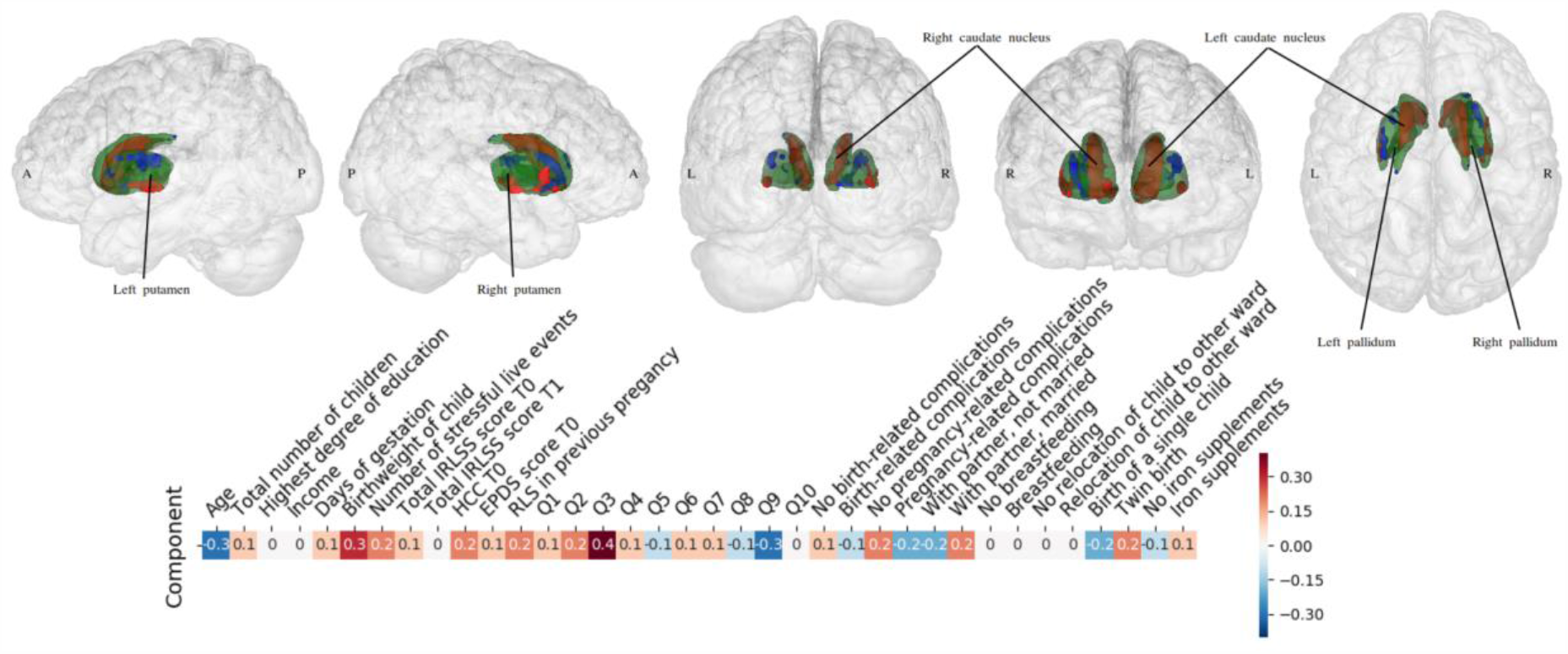
Specific patterns linking the basal ganglia to the phenotypical predictors of RLS. *IRLSS score T0 = Severity of RLS symptoms in pregnancy; IRLSS score T1 = Severity of RLS symptoms 12 weeks postpartum A targeted post-hoc CCA was applied to find coherent associations between the basal ganglia volume changes and changes in the patients’ anamnestic and clinical indicators. While the above-mentioned analysis (Fig. 2) considered the whole brain as measured by volume estimates, the present analysis focused on the left and right basal ganglia at a more fine-grained voxel resolution. Green indicates the anatomy of the target regions as part of the basal ganglia. In the top row, the loadings of the basal ganglia voxels are plotted from the left, right, posterior, anterior and superior views. The green area contours expose the outer shape of the basal ganglia voxels, which cover the striatum (putamen and caudate nucleus) and the pallidum. Within the basal ganglia, the blue area indicates negative CCA loadings associated with those specific voxels while the red indicates positive CCA loadings for those specific voxels. The bottom row depicts the loadings of the patient’s behavioral and clinical indicators (blue for negative and red for positive). While having a baby with higher birth weight and having the unpleasant sensations in the legs or arms alleviated by exercise (Q3) were strongly linked to an increased size of the body and head of the left and right caudate nuclei and the left and right inferior putamen, being young and not being so affected by RLS (Q9) were found to be associated with a decreased size of the superior part of the left and right putamen. In sum, our analysis revealed strong associations between anamnestic changes and changes in the medial and lateral striatum within the basal ganglia.

In short, major changes were found in the basal ganglia, with an inter-hemispheric increase in the size of the caudate and the inferior part of the putamen as the size of the baby and RLS-related sleep deprivation increased.

## DISCUSSION

In a large neurological-obstetrical sample of 308 postpartum women, participants with pregnancy-related RLS were identified and compared with the rest of the sample. Furthermore, a number of clinical and anamnestic indicators of RLS in pregnancy were assessed by leveraging state-of-the-art machine learning algorithms. Within the group of women with pregnancy-related RLS, the phenotypical predictors of the condition were assessed by means of a brain structure analysis of the brain phenotype model.

The RLS prevalence rate in our sample was found to be 19%. While in 30% of the cases the symptoms were still reported 12 weeks postpartum, 70% of the women experienced a complete recovery of RLS symptoms shortly after delivery. The majority of participants experienced the RLS symptoms as most severe and most frequent in the last trimester of pregnancy. In line with previous findings (Innes et al., 2015; Ramirez et al., 2013), those afflicted by RLS in our sample were older and more often affected by gestational diabetes than their unaffected counterparts. We found also that women experiencing RLS in pregnancy reported to experience SLE more often than those without RLS (Chandan et al., 2019). Additionally, there were group differences in terms of marital status, with women in the RLS group being more often not married.

Examining the possible predictive factors of RLS in pregnancy, we found a history of RLS to be the strongest predictor of the condition in current pregnancy, which is in line with previous reports (Cesnik et al., 2010). In IRLSS, items describing the degree of urge to move the legs and the degree of RLS symptom relief afterword were better predictors of pregnancy-related RLS symptoms compared to those describing daily activity, mood or sleep.

As regards the relationship between whole-brain gray matter morphology and phenotypical predictors, high HCC, being married and receiving iron supplements were found to be associated with increased volumes of the left superior temporal pole, the left inferior frontal gyrus and the basal ganglia (the right and left caudate nucleus and putamen). Previous studies have foreshadowed a possible physiological link between factors such as the circadian rhythm of cortisol secretion or iron deficiencies in the central nervous system (CNS) and manifestation of idiopathic RLS. In particular, an evening and early night hour RLS symptom increase has been suggested to be moderated by low-dose hydrocortisone (Hornyak et al., 2008; Rizzo and Plazzi, 2018). Apart from the circadian rhythm of cortisol secretion, reduction of brain iron is thought to be one of the risk factors in the development of RLS symptoms (Rizzo and Plazzi, 2018), the iron deficiency being attributed to dysregulations in the dopamine system (Earley et al., 2014). Also noteworthy is the relationship seen between marital status and increased basal ganglia structures in RLS patients. Married individuals are suggested to be healthier (Robards et al. 2012; Svetel et al. 2015) and therefore less likely to be exposed to stressors compared to those who either never married or were previously married (Chin et al., 2017). As also suggested by previous research, the positive relationship between brain volume in the striatum and frontal areas and such factors as higher HCC, iron supplementation or being married are likely associated with more favorable prognoses with respect to pregnancy-related RLS. At the same time, a history of RLS, not being married and not receiving any iron supplement were found to be associated with a decreased volume size of the (right inferior) parietal lobule, the posterior cingulate, and the right medial orbitofrontal and frontal (left superior frontal gyrus) cortices. The same negative relationship of brain volume in the above-mentioned regions was seen with the severity of RLS-related sensations, the intensity of the urge to move the legs and the number of SLE. Based on these observations, we suggest that the association between reduced brain volume and factors such as history of RLS, not being married, higher number of SLE or being more strongly affected by RLS symptoms is probably indicative of a less favorable prognosis for pregnancy-related RLS. This assumption is supported by the fact that a history of RLS and the intensity of the urge to move the legs were found to be the strongest predictors of pregnancy-related RLS symptoms in our sample. Previous research has indicated links between SLE such as child maltreatment and the development of a set of somatic and visceral central sensitivity syndromes like chronic pain, irritable bowel syndrome as well as RLS (Chandan et al., 2019).

Taken together, our results suggest that pregnancy-related RLS (similarly to idiopathic RLS (Margariti et al., 2012; Rizzo and Plazzi, 2018)) is linked to the striatal and frontoparietal structures. As the whole-brain results of our study show, the most important and interhemispheric variations occur in the basal ganglia, with the medial and lateral striata contributing the most to the clinical and anamnestic differences in the pregnancy-related RLS group. Along with our results, the human lesion studies (Lee et al., 2009; Shiina et al., 2019; Woo et al., 2017) as well as the recent RLS animal model studies (Guo et al., 2017) underscore the crucial role of the basal ganglia in the manifestation of the RLS spectrum. Based on the brain phenotype model, we conclude therefore that different factors such as stress response, history of SLE or life circumstances (e.g. marital status) or iron deficiencies in the CNS contribute to the manifestation of RLS in pregnancy. Interconnected in a complex way, these factors, along with genetic predisposition, likely lead to the dysfunction of the mesolimbic and nigrostriatal dopaminergic pathways, triggering abnormalities in the limbic/nociceptive and sensorimotor networks (Rizzo and Plazzi, 2018) and the development of RLS symptoms. The investigation of RLS in pregnancy in a brain phenotype model has the potential of helping augment our understanding of the heterogeneity of the spectrum.

## Data Availability

The data are available on request from the corresponding author.

## Acknowledgments

This study was funded by the rotation program of the medical faculty of University Hospital RWTH Aachen and the Deutsche Forschungsgemeinschaft (DFG, CH 1718/2-1) and was supported by the International Research Training Group (IRTG 2150) of the German Research Foundation (DFG).

DB was supported by the Healthy Brains Healthy Lives initiative (Canada First Research Excellence fund), Google (Research Award), and by the CIFAR Artificial Intelligence Chairs program (Canada Institute for Advanced Research).

## Author Contribution

Conceptualization, N.C., D.B. and T.G; Methodology, D.B.; Software, D.B.; Validation, D.B., N.C. and J.L.; Formal Analysis, J.L. and N.C; Investigation, M.F., S.S, P.S, and T.G; Resources, N.C. and T.G; Data Curation, S.S., and P.S.; Writing – Original Draft Preparation, N.C.; Writing – Review & Editing, N.C., JL., TG., MF., P.S., S.S, and DB; Visualization, J.L.; Supervision, D.B.; Project Administration, M.F.; Funding Acquisition, N.C.

